# Molecular characterisation of longitudinally collected circulating cell-free DNA in HPV+ve and HPV-ve oropharyngeal cancer

**DOI:** 10.1101/2020.09.07.20189704

**Authors:** John P Thomson, Sophie J Warlow, Martyna Adamowicz, Helen Thain, Kate Cuschieri, Lucy Q Li, Brendan Conn, Ashley Hay, Iain J Nixon, Timothy J Aitman

## Abstract

Oropharyngeal squamous cell carcinoma (OPSCC) is an increasing global health problem and is divided into two types dependent on association with human papillomavirus (HPV), with a more favourable prognosis in virus-associated tumours. Current methods of establishing viral aetiology, assessing response to therapy and clinical monitoring rest on tissue biopsy, clinical examination and post-treatment imaging. However, tissue biopsy is invasive and carries significant risk of morbidity, and post-treatment scans are frequently indeterminate. Analysis of cell-free DNA (cfDNA) from the circulation provides a minimally invasive method for detecting and monitoring cancer-derived DNA fragments, with the potential for enhancing clinical care. Through the longitudinal collection of 166 blood samples in 67 OPSCC patients we evaluate the utility of three cfDNA analysis methods: droplet digital PCR (ddPCR) and fragment size analysis in both HPV+ve and HPV-ve disease, and ultra-deep sequencing in patients with HPV-ve disease. We show that ddPCR analysis of cfDNA for five HPV types (16, 18, 31, 33 & 35) is strongly concordant with existing clinical assays (p16 immunohistochemistry (IHC) and quantitative PCR analysis of solid tumour tissue) and that cfDNA fragment size was reduced in OPSCC patients compared to healthy controls. Sequential ddPCR measurements of cfDNA HPV copy number showed a decrease to undetectable levels in all 30 HPV+ve patients in at least one of their post-treatment samples and a corresponding increase in cfDNA fragment size in patients who had a complete response to chemoradiotherapy. In two HPV+ve patients, clinical decision-making based on HPV ddPCR of cfDNA may have led to earlier detection of relapse in one patient or avoided surgical exploration in a second patient, which led to resection of tissue that did not harbour malignancy. In HPV-ve disease, ultra-deep sequencing identified tumour-derived somatic mutations of circulating cfDNA in genes such as TP53 and members of the ERBB family that are potential markers of therapeutic responsiveness and patient prognosis. Together our data suggest that analysis of circulating cfDNA can enhance current clinical strategies for assessing therapeutic response and disease monitoring in both HPV+ve and HPV-ve OPSCC.

## Introduction

Oropharyngeal squamous cell carcinoma (OPSCC) is an increasingly prevalent cancer, linked in part to tobacco smoking, alcohol consumption and association with human papillomavirus (HPV)[1]. HPV infection, particularly type 16 (HPV16), is now recognized as a major cause of OPSCC. HPV associated (HPV+ve) OPSCC has significantly different clinical, radiological and prognostic characteristics from typical OPSCC with no viral association (HPV-ve) [2, 3]. The better prognosis and potential differences in therapeutic approaches in HPV+ve OPSCC underline the importance of accurate identification of HPV status in patients with this condition [4].

Current pre-treatment assessment of OPSCC includes clinical examination and cross-sectional imaging with targeted biopsy and histopathological diagnosis supported by immunohistochemistry(IHC) against the protein p16 and/or PCR testing of biopsy material for HPV [3]. This is followed by chemoradiotherapy (CRT) in most cases, with 12-week post-treatment imaging to gauge response to therapy and need for salvage surgery. Further follow-up entails regular clinical assessment with imaging and biopsy where required. Biopsy of post-treatment OPSCC can be challenging however, and even with recent advances in cross-sectional and functional imaging, a significant number of patients have indeterminate results [5, 6]. Such patients must then elect either for continued surveillance with repeated imaging and long periods of uncertainty, at significant financial and psychological cost, or further treatment often in the form of neck dissection. Because of the poor predictive power of post-treatment imaging, particularly in HPV+ve disease, surgical procedures can result in resection of tissue in which no active disease is detected. Such procedures remain associated with significant morbidity, often long-term. A more reliable marker of residual or recurrent disease would therefore have considerable benefits for patient management.

The analysis of circulating tumour-derived DNA (ctDNA) from patient blood, or “liquid biopsy”, represents a minimally invasive approach to diagnosis and management, with potential advantages for clinical care of patients with head and neck cancers [7]. There has been significant interest in developing liquid biopsy approaches in many cancer types including HPV+ve OPSCC, for monitoring treatment response, predicting relapse and providing early detection of tumour recurrence [8–11]. In certain cancers, detection of actionable tumour-derived somatic mutation from ctDNA has already been used or proposed as an indication for targeted therapy [12–14].

The association of the majority of cases of OPSCC with HPV infection offers an opportunity for diagnosis and disease tracking of HPV+ve cases through detection of HPV in plasma cfDNA and a number of studies have taken this approach, detecting the presence of several HPV types by ddPCR or amplicon-based sequencing [11, 15–19]. However, no such liquid biopsy markers are available for HPV-ve disease, for which disease monitoring currently relies on clinical examination, imaging, surgical biopsy and histopathological analysis. In contrast to genome analyses of solid tumour material, sequencing of cfDNA to detect tumour-derived somatic mutations is challenging, largely due to the low fraction of tumour-derived DNA in a background of non-tumour DNA present in plasma [20, 21].

Here we evaluate the role of liquid biopsy assessment across all patients with OPSCC. In order to study both HPV+ and HPV-ve disease, two different approaches were developed. In HPV+ve disease, we use HPV ddPCR of cfDNA to define HPV status in comparison to conventional p16 immunohistochemistry and qualitative PCR (PCR) of solid tumour biopsies and carry out sequential sampling to correlate HPV cfDNA copy number with therapeutic response and clinical course. In HPV-ve disease, we carry out ultra-deep targeted sequencing of cfDNA using a protocol that includes bait enrichment, unique molecular identifiers (UMIs) and error suppression informatics, as a proof of concept of this approach to robustly detect somatic mutations in plasma cfDNA of HPV-ve OPSCC patients, and as a means of advancing the route to molecular classification and monitoring of this poor prognosis form of OPSCC.

## Materials and methods

Patients were screened and approached following discussion at the South East Scotland Multidisciplinary Head and Neck Cancer Meeting, with samples collected under the Lothian NHS Research Scotland BioResource and ethics approval by the East of Scotland Research Ethics Committee (Reference: SR1171). 47 male and 20 female consecutive patients were consented during their pre-treatment work-up with 100% of those approached agreeing to consent. Patients had a median age of 60 years (min 38, max 86). The majority of patients (46/67) received combined CRT, with further details of treatment regimens as shown in Table 1.

**Table 1.** Patient demographics, tumour stage and treatment

|  |  |  |  |  |
| --- | --- | --- | --- | --- |
| **Age (Median (range))** |  | 50 (35-86) |  |  |
| **Gender (Male/Female)** |  | 47/20 |  |  |
| **Subsite** | Tonsil | 39 |  |  |
|  | Base of tongue | 22 |  |  |
|  | Other | 6 |  |  |
| **P16 (+ve/-ve)** |  | 52/15 |  |  |
| **TNM Stage** |  | **T** | **N** | **M** |
|  | 0 | 3 | 11 | 67 |
|  | 1 | 23 | 10 |  |
|  | 2 | 14 | 46 |  |
|  | 3 | 5 | 0 |  |
|  | 4 | 22 |  |  |
| **AJCC STAGE 8** |  |  |  |  |
|  | 1 | 35 |  |  |
|  | 2 | 9 |  |  |
|  | 3 | 14 |  |  |
|  | 4 | 9 |  |  |
| **Treatment** |  |  |  |  |
|  | RT | 6 |  |  |
|  | CRT | 46 |  |  |
|  | Induction chemo + CRT | 2 |  |  |
|  | CRT + immunotherapy | 2 |  |  |
|  | Surgery | 2 |  |  |
|  | Surgery + RT | 3 |  |  |
|  | Surgery + CRT | 2 |  |  |
|  | Palliative CRT | 4 |  |  |

Patients were categorised as having progressive disease or a complete or partial response by 12-week post-treatment contrast-enhanced CT scan. Those patients with necrotic, contrast enhancing or size-significant (>1cm) lymphadenopathy and patients with radiological evidence of residual primary disease following therapy were considered suspicious for persistent disease. For the purposes of this study the results of cross sectional (CT) and functional CT-FDG PET were combined to characterise patients as having a complete response, partial response or those with progressive disease. Patients with a complete response entered clinical follow up. Patients who had a partial response, with “high-grade uptake” on the CT-FDG PET and who were fit for additional treatment underwent salvage neck dissection. Patients who had a partial response with “low-grade uptake” on CT-FDG PET were considered on an individual basis, including clinical, tumour and functional characteristics and were offered either salvage surgery or continued radiological surveillance after the 12 week assessment point. AJCC staging (8^th^ edition) was recorded to define overall disease stage [22]. Patients were grouped as either “non-”, “current-” or “ex”-smokers and into one of four alcohol consumption groups: “non / minimal”, “occasional” (< = 10 units per week (U per/week)), “moderate” (11–19 U per/week), “excess” (> = 20 u per/week).

### Collection and analysis of HPV status in solid tumour tissue

All patients underwent per oral biopsy of primary and/or ultrasound-guided biopsy of nodal tissue. Sections of formalin-fixed paraffin-embedded (FFPE) blocks were selected and reviewed by a pathologist with a special interest in head and neck cancer for histological diagnosis. Immunohistochemistry (IHC) for p16 was carried out on 3μm sections using the CINtec p16 Histology assay (Roche Laboratories) for qualitative detection of the p16INK4a protein on FFPE tissue sections, using a Leica Bond III automated immunostainer. IHC for p16 was assessed by the pathologist and considered positive if there was strong diffuse nuclear and cytoplasmic staining present in > 70% of malignant cells [23]. Qualitative PCR (PCR) HPV typing was performed on DNA extracted from a 10 micron section of the relevant block as described [24], by PCR amplification of L1 sequences of 24 HPV types (HPV 6/ 11/ 16/ 18/ 26/ 31/ 33/ 35/ 39/ 42/ 43/ 44/ 45/ 51/ 52/ 53/ 56/ 58/ 59/ 66/ 68/ 70/ 73 & 82) followed by genotyping on the Luminex 200 platform (R&D Systems).

### Collection of blood and cfDNA extraction

Pre-treatment blood samples were collected from all patients on recruitment to the study. Although some clinic visits were cancelled due to Covid-19 lockdown, where possible post-treatment blood samples were also collected from each patient. Total blood collections numbered 166. The median follow-up time was 15 weeks post-completion of treatment, ranging from 3-55 weeks. Whole blood was collected into PAXgene® Blood ccfDNA Tubes (Qiagen, #768115) and stored at 4°C before processing within 48 hours. Plasma was separated by centrifugation at 1000g for 10 minutes at 18°C and then at 13,800g for 5 minutes at 18°C, prior to transfer into 1.8ml cryotubes (Nunc, #10004220) for storage at −80°C until the time of cfDNA extraction. Human K2 EDTA control plasma was purchased from BioIVT Seralabs (Detroit, USA).

For cfDNA size and concentration fragment analysis and for ddPCR, cfDNA was extracted from 0.5-1ml of plasma using the QIAamp MinElute Virus Spin Kit (Qiagen, #57704) with modifications to the manufacturer’s protocol to improve enrichment (see Supplementary Material and Methods). For genomic sequencing on the Illumina TSO500 platform, cfDNA was extracted from paired pre- and post-treatment plasma (2-5 mls), using the QIAamp Circulating Nucleic Acid Kit (Qiagen, #55114), with modifications to improve enrichment (see Supplementary Material and Methods). cfDNA was stored at −20°C until required for ddPCR or library preparation for genomic sequencing.

### HPV ddPCR

ddPCR assays were carried out to detect the 5 most prevalent HPV genotypes using primers designed and developed by Jeannot et al (HPV16 & 18) and Chera et al (HPV31, 33 & 35) [15, 18]. Further detail is given in Supplementary Materials and Methods. Assays were optimised and performed in accordance with the Minimum Information for Publication of Quantitative Digital PCR Experiments (dMIQE) guidelines [25] (Supplementary Tables 1–3). cfDNA samples were assayed in 10 replicates, using the total available volume of cfDNA after QC. Positive, negative and no template controls (NTC) were included in every ddPCR run. ddPCR data was analysed using fluorescence amplitude data for individual droplets extracted by the QuantaSoft software v.1.7.4.0917 (Bio-Rad). Resulting .csv files were analysed using the open-access web-based JavaScript program *definetherain* (https://github.com/jacobhurst/definetherain) [26], which uses the k-means algorithm to cluster the negative and positive populations within a control sample. Further details are given in Supplementary Materials and Methods.

### Analysis of cfDNA fragment size and concentration

Where sufficient sample was available following ddPCR assay, cfDNA samples were characterised by electrophoresis using the Cell-free DNA ScreenTape Assay on the TapeStation 4200 platform (Agilent). TapeStation Analysis Software v3.2 (Agilent) was used to calculate QC metrics, including average fragment size (average size in bp in window 50bp-700bp), peak size of the mononucleosomal fragment and overall cfDNA concentration (pg/μl). Statistical analyses and resulting plots were performed using R (version 3.5.1). Statistical significance between groups was calculated by 1-tailed Student’s t-test.

### cfDNA sequencing on the Illumina TSO500 cfDNA platform

cfDNA libraries were prepared according to the TruSight Oncology 500 ctDNA Kit (Illumina, #20039252) manufacturer’s instructions with an adjustment of input cfDNA based on available plasma (4ng-30ng cfDNA). Seraseq ctDNA Complete Mutation Mix AF 0.5% (30ng, Seracare, #0710-0531) was used as a positive control. For details of library preparations please refer to supplementary methods. Library pools were sequenced on the NovaSeq 6000, using two S2 flowcells (Illumina, #20012860) to generate 2x 150bp paired-end reads with dual indexing. Across the 1.94 Mb targeted regions, ultra-deep sequencing returned an average 1,151,350,985 reads, min: 969,250,914, max: 1,303,460,528.

### Sequence analysis

Analysis was conducted using the Illumina TruSight™ Oncology 500 ctDNA Local App on the Illumina DRAGEN server. Data processing generated a variant call file (*.vcf) for small variants and reports of mutations per megabase scores for tumour mutational burden (TMB) and copy number variation (CNV). Further details are given in Supplementary Materials and Methods. For downstream analysis, VCF files were further filtered to remove synonymous and intronic variants as well as variants with variant allele frequencies > 20%. Post-filtering analysis yielded a mean exonic coverage of 1026.9X (min 285X, max 1502X). Supervised mutational analysis, such as oncoplot analyses, were visualised using the R package maftools [27].

## Results

### Patient characteristics

Age, gender, primary tumour sub-site, TNM and AJCC8 stage details are shown in Table 1. Of the 67 patients recruited, 39 (57%) had primary tonsillar disease, 22 (32%) base of tongue and the remaining 6 (11%) had disease at other sites (unknown primary, soft palate and lateral pharyngeal wall) (Figure 1), which is representative of the expected demographic in the UK [28]. Four patients were treated with palliative intent (6%). Of those patients treated with curative intent, 7 patients (10%) had surgery with or without adjuvant therapy. The remaining 56 patients were treated with a radiation-based approach (concurrent chemoradiation in 46 (69%), radiation alone in 6 (9%), induction chemotherapy followed by concurrent chemoradiation in 2 (3%) and chemoradiation plus adjuvant immunotherapy in 2 (3%)).

**Figure 1.**
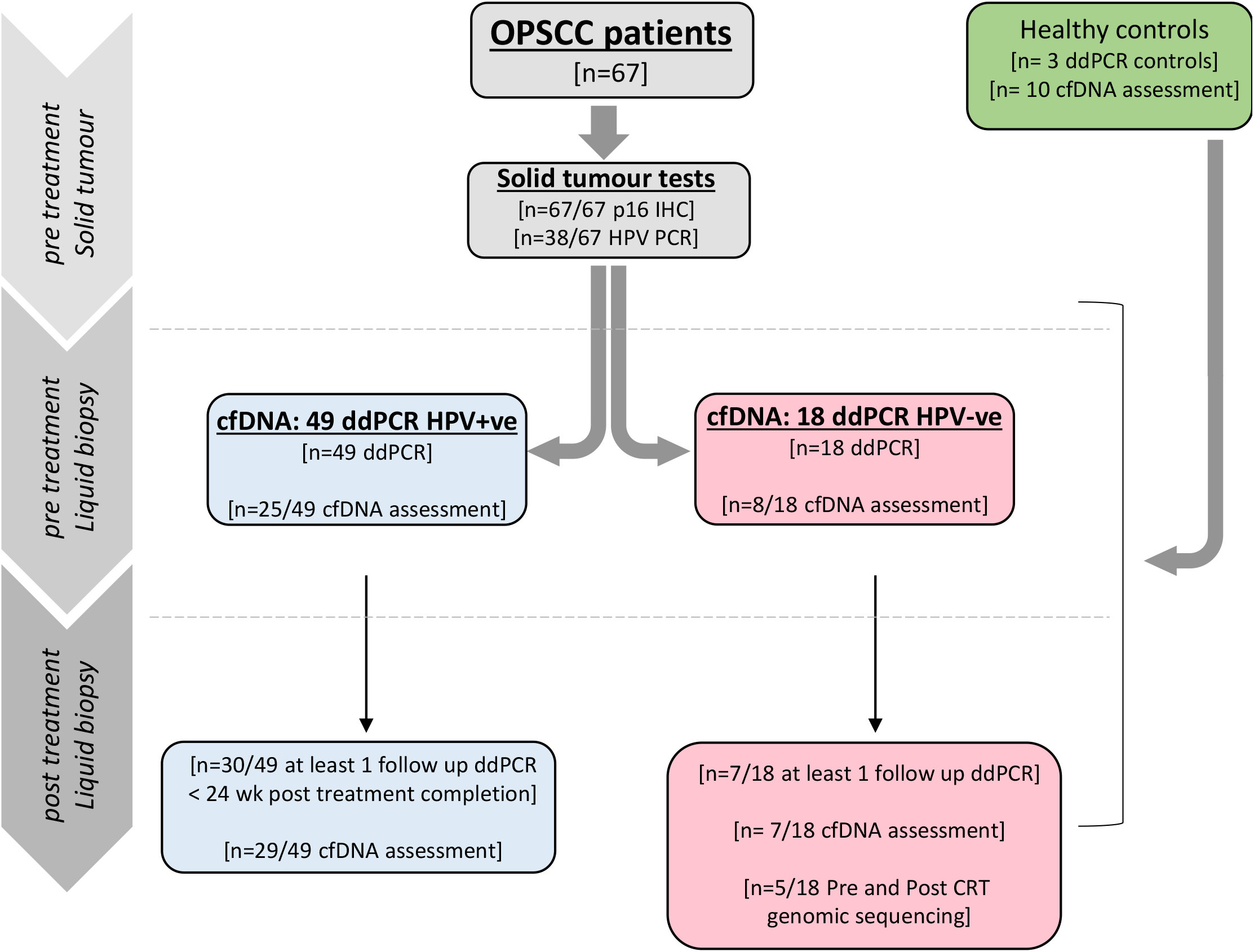
Flow diagram of patients and samples in the study. Workflow describes the number of patients recruited to the study as well as the number included in each test.

### Comparative analysis of cfDNA HPV ddPCR and tumour-based classification

Conventional assessment of HPV status in OPSCC patients by p16 IHC or HPV PCR of tumour DNA requires access to solid tumour tissue. We sought to benchmark HPV analysis of plasma cfDNA, obtained from a simple blood draw, against p16 IHC and HPV PCR from solid tumour. We therefore established ddPCR assays of cfDNA for the five most prevalent HPV types (HPV 16, 18, 31, 33 & 35) and measured HPV copies present in plasma cfDNA in the 67 recruited OPSCC patients (Figures 1, 2; Supplementary Figure S1). Each primer and probe set was specific to its HPV genotype (Supplementary Figures S2, S3).

**Figure 2.**
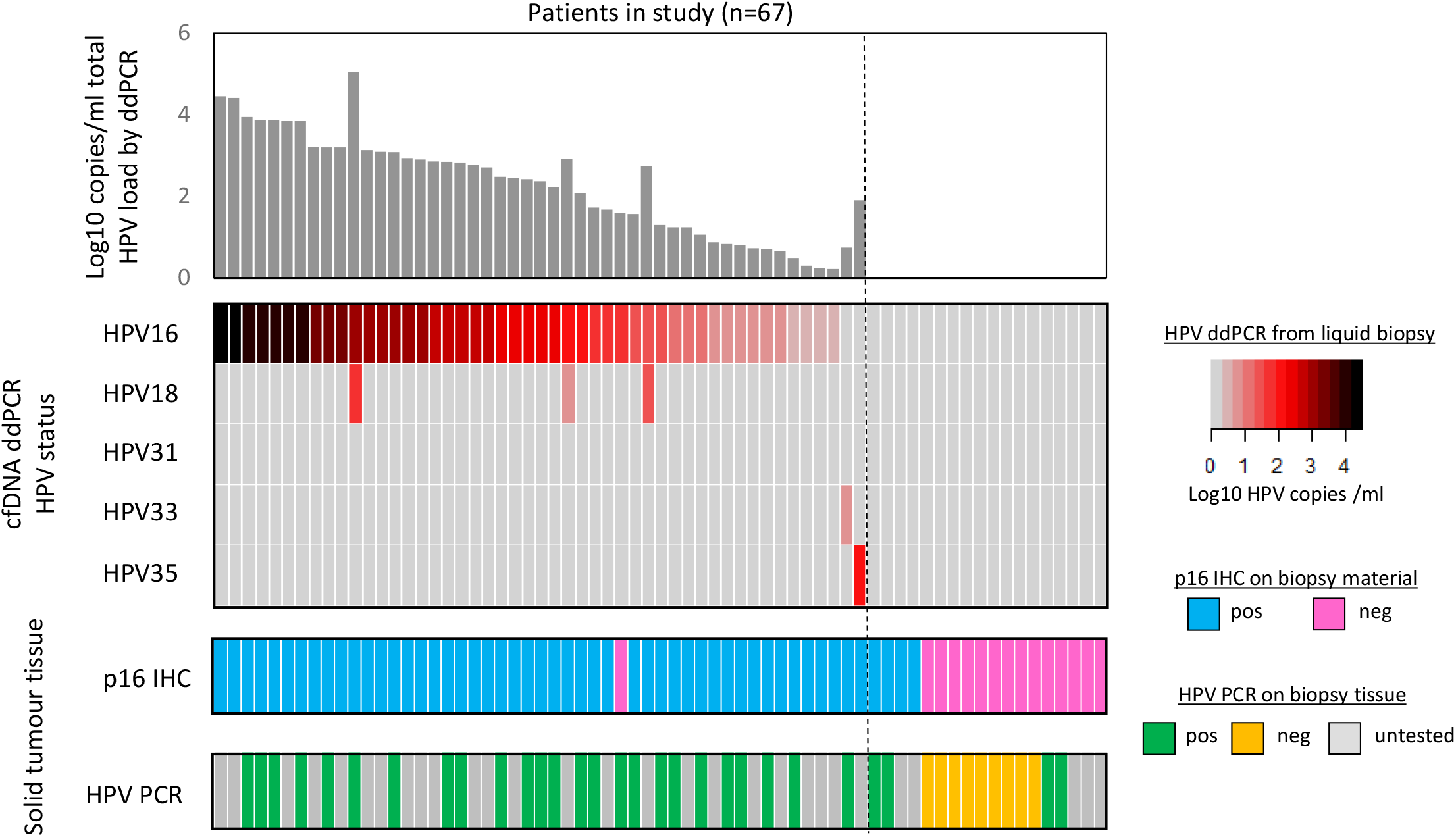
Comparisons between blood based ddPCR assay and solid tumour assays. **A.** Heatmap of copies of HPV in the blood (plotted as Log10 copies/ml plasma) for the five most prominent HPV genotypes (rows). Each column denotes a patient. Dark red: high levels HPV, light red: low levels HPV. Grey bars denote samples with no detectable copies beyond background levels. Bar plot above displays the combined copies/ml for all genotypes in each patient. Plots below describe results from matched solid tumour biopsy material for both p16 immunohistochemistry (IHC). Blue bars: p16 positive, pink bars: p16 negative) and HPV PCR (green bar: HPV positive, yellow bar: HPV negative, Grey bar: no data available). **B.** Boxplots of log10 copies of HPV/ml as detected by ddPCR against i) smoking status, ii) alcohol consumption, iii) AJCC stage 8 and iv) site of the primary tumour.

We detected HPV by ddPCR in plasma cfDNA of 49 of the 67 patients, with a wide range of copy numbers (median 279 copies/ml; range 1-30,380 copies/ml; Figure 2; Supplementary Figure S3). The overwhelming majority of HPV+ve patients (47/49 patients) were positive for HPV16 (median 297 copies/ml). One patient was positive for HPV33 (5 copies/ml) and one patient for HPV35 (84 copies/ml). Three patients were ddPCR positive for both HPV18 (median 154 copies/ml) and HPV16 (Figure 2).

Next, we compared the results of cfDNA ddPCR to matched solid tissue biopsy tests for p16 IHC and HPV PCR. 48/49 cases classified as HPV+ve by cfDNA ddPCR stained positively for p16 in matched tumour tissue (98% concordance), whilst 14/18 cases classified as HPV-ve by cfDNA ddPCR were also negative for p16 IHC (78% concordance) giving an overall concordance of 93% between cfDNA ddPCR and p16 IHC results (Figure 2). Where sufficient tumour DNA was available (n = 38), we also carried out qualitative PCR and genotyping, which detects 24 HPV types, including all established high-risk types, on DNA extracted from solid tumour tissue. HPV PCR results agreed with p16 IHC data in 35/38 (92%) of matched tumour samples, whereas HPV PCR results agreed with cfDNA ddPCR results in 34/38 (89%), although two of the samples showing discordance between cfDNA ddPCR and tumour PCR were concordant between cfDNA ddPCR and p16 IHC (Figure 2).

Overall, in patients who were tested by all three assays, 100% (49/49) of cfDNA ddPCR HPV+ve results were also positive by either IHC or PCR. Of the 13 cfDNA ddPCR HPV-ve patients who were tested by all three assays, 85% (11/13) were also negative by either p16 IHC or HPV PCR. Taken together, the data suggests that blood-based ddPCR results are strongly concordant with existing clinical assessment methods employed on solid tumour tissue.

### cfDNA fragment length and concentration

Recent studies have shown that differences in cfDNA fragment length and concentration can be exploited to enhance sensitivity for detecting the presence of ctDNA across a range of cancer types[29]. Where sufficient plasma was available following ddPCR assessment (33/67 of pre-treatment samples), we carried out electrophoresis of cfDNA in pre-treatment patient samples. No significant differences in cfDNA fragment size or concentration were seen between patients with HPV+ve and HPV-ve disease (Supplementary Figure S4). However, average fragment size and mononucleosomal peak size were significantly higher in healthy controls than in the set of all OPSCC patients in whom fragment size data was available (Figure 3A; P = 1.69 E-11 and P = 0.003 respectively). Fragment size was also higher in patients with primary tonsillar tumours than primary tumours in the base of the tongue (Figure 3B; P = 0.01). Fragment sizes did not differ when analysed by tumour stage, smoking status or alcohol consumption (Figure 3C-E). cfDNA concentration did not differ between healthy controls and OPSCC patients or between OPSCC patients analysed by stage, sub-site of primary tumour, smoking status or alcohol consumption (Supplementary Figure S5).

**Figure 3.**
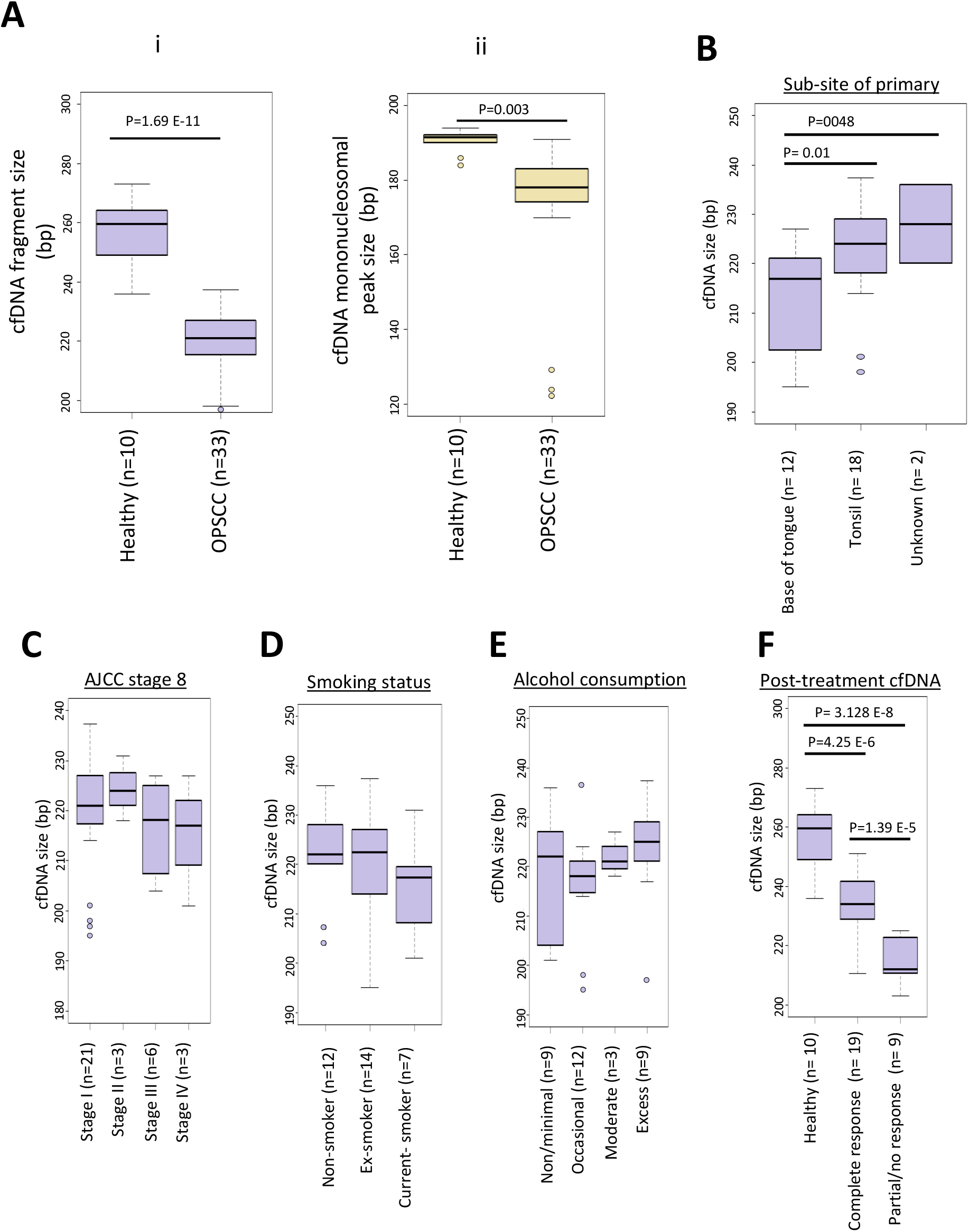
cfDNA fragment size as a general biomarker of OPSCC. **A. (i)** Box plot of average fragment size, in range 50–700bp, between OPSCC patients and healthy controls. **(ii)** Box plot of mono nucleosomal peak size between OPSCC patients and healthy controls **B-E.** cfDNA fragment sizes categorised by sub-site of the primary tumour **(B)**, AJCC stage 8 criteria **(C)**, smoking status **(D)** and alcohol consumption **(E). F.** Post-treatment OPSCC bloods by outcome of 12 wk PET/CT scans relative to healthy control samples. Boxes represent 1st to 3rd quartile, with the median labelled as the central line. Boxplot whiskers extend to the largest value within Q3 + 1.5 times interquartile range P values are shown where P< 0.05 by 1-tailed students T-Test. In Panel B, data was unavailable for one patient with tumour located in the soft palate.

### Longitudinal monitoring of HPV+ve OPSCC patients

To monitor treatment success in OPSCC patients longitudinally, we collected sequential post-treatment blood samples in 30 of the 49 HPV+ve and in 7 of the 18 HPV-ve patients, at all scheduled clinic visits. We observed a decrease to zero HPV copies/ml in cfDNA of all HPV+ve patients in at least one of their post-treatment samples, regardless of the starting HPV levels (Figure 4A). HPV-ve OPSCC patients displayed consistently negative results across all time points reiterating the specificity of the ddPCR assays across time points and treatment stage (Supplementary Figure S6). All but two of the 30 HPV+ve patients with follow-up blood samples (93.3%) had undetectable HPV levels in their first post-treatment blood draw (average 9 weeks, range 2-21.5 weeks). Of the remaining two, whom we designate as “slow response” patients, one had undetectable levels by 14 weeks post treatment (Figure 4A). The second slow response patient (Patient #04) was unique in that this was the only HPV positive case treated with combined CRT and adjuvant immunotherapy (Supplementary Figure S7). In this case, HPV levels remained high 18 weeks post-treatment, though HPV levels ultimately fell to undetectable levels at 35 weeks post-treatment. The patient made a complete clinical and radiological response to treatment.

**Figure 4.**
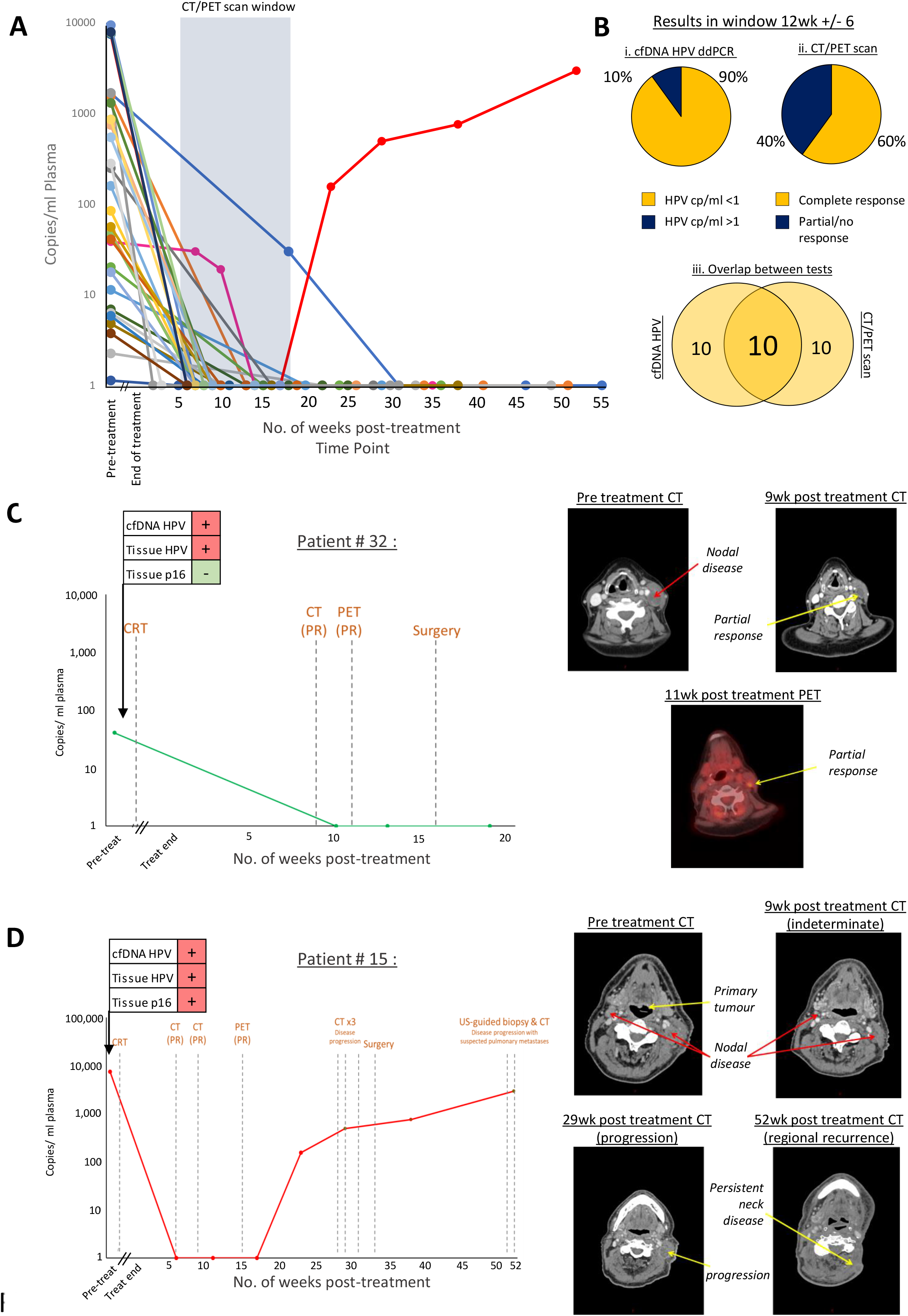
Longitudinal monitoring of HPV Copies/ml in plasma in HPV+ve patients. **A.** Plot of the levels of HPV over time (copies/ml) prior to and following completion of treatment for patients with follow up bloods (n = 30). Data plotted as pre-treatment load and then at number of weeks following end of treatment. Blue box denotes window with overlap to 12 week CT/PET scan (12wk treatment completion +/− 6 wks). **B.** Pie charts plotting the i) number of HPV ddPCR null and positive samples and ii) combined CT/PET results within analysis window (blue box figure 4A). Percentages are shown next to each plot. Venn diagram displaying the overlap between results are shown below in iii). **C.** Case study for the single patient who had discordant solid tissue results and **D** the single patient in whom relapse occurred. For both C and D, copies of HPV detected in cfDNA with clinical milestones overlaid (grey dashed lines) are shown on the left. “PR”: partial response. For each case, CT and PET scans from select time points are shown on the right. Suspected nodal disease indicated by red arrows. Suspected tumour indicated by yellow arrows.

Next we compared the results of the ddPCR tests in HPV+ve patients to those of routine imaging. As post-treatment contrast-enhanced CT is typically performed at around 12 weeks after completion of treatment we restricted our analysis to the 20 patients from whom blood samples were collected within a similar time window (12 +/− 6 weeks, blue box in Figure 4A). 12 of the 20 patients (60%) were classified as showing complete response by imaging (CT +/− PET) with 8 patients showing only partial or no response. In contrast, 18 of the 20 patients (90%) were classified as having no detectable HPV levels by cfDNA ddPCR, suggesting a complete response in these patients.

To test if cfDNA fragment size or concentration were indicative of therapeutic response across patients with a post-treatment blood collection, we quantified these parameters in the first available sample following the end of therapy. cfDNA fragment size was significantly higher in patients who showed a complete response than in those with a partial or no response on imaging (P = 1.39 E-5), although fragment sizes in both groups were still lower than those in healthy controls (P < 10 E-5 for both comparisons) (Figure 3F). No differences were seen in post-treatment cfDNA concentrations between these groups (Supplementary Figure S5F).

### cfDNA HPV tests can guide in cases of discordant biopsy results

Current clinical assessment of HPV status in OPSCC is based on an agreement in both p16 IHC and HPV specific tests from solid tissue material. In our cohort, one case (Patient #32) had a biopsy that was negative for p16 IHC but was positive by HPV PCR and ddPCR of cfDNA. Longitudinal assessment of cfDNA HPV copy number in this patient revealed a drop from 40.82 copies/ml pre-treatment, to undetectable HPV copy number in the first post treatment sample 10 weeks after completion of CRT, persisting as undetectable in all further samples (Figure 4C). Based on the negative p16 IHC result, the patient’s tumour was classified as HPV-ve and because repeat CT scans and FDG-PET scan indicated a partial response or possible disease progression, with low-grade FDG uptake on the PET scan, salvage surgery was undertaken at 33 weeks after the initial treatment. Salvage surgery resulted in a hypoglossal nerve injury and pathological analysis of surgically removed tissue showed no detectable tumour, consistent with the undetectable levels of HPV by ddPCR in post-treatment cfDNA after a previous positive pre-treatment result. Management based on combined cfDNA HPV ddPCR and solid tissue HPV PCR would have classified this patient as HPV+ve with a reduction in cfDNA HPV by ddPCR following CRT to undetectable levels, which may have suggested that surgical intervention was not required.

### cfDNA monitoring can provide an early indication of relapse

In 29 of the 30 HPV+ve OPSCC patients with follow up-bloods, once HPV cfDNA levels had fallen to undetectable levels, HPV levels remained undetectable for the remainder of the study (Figure 4A). One case (Patient #15), however, displayed a marked rise in HPV cfDNA copy number at 23 weeks and subsequently at 29 weeks after treatment with CRT, having previously shown zero copies at 6 and 11 weeks post-treatment. (Figure 4D). This patient had presented with stage IV OPSCC located at the base of the tongue with bilateral neck disease and was classified as HPV+ve by solid tissue p16 IHC and PCR, and by ddPCR of cfDNA. Clinical assessment by combined CT and PET scanning indicated only a partial response to CRT with low-grade uptake on CT-FDG PET which was considered an indeterminate result on multi-disciplinary team assessment. Due to advanced age, bilateral neck disease and uncertainty of disease response on imaging, the decision was made to monitor the patient with serial CT scans. The disease progression suggested by the rise in HPV copy number at week 23 was confirmed 9 weeks later by CT scan, leading to surgical intervention at week 33 (Figure 4D). cfDNA HPV levels increased further after surgery (492 HPV copies/ml 4 weeks prior to surgery vs. 753 HPV copies/ml and 2,939 copies/ml 5 weeks and 19 weeks respectively post-surgery). Disease progression was confirmed on tissue biopsy and the patient was commenced on palliative immunotherapy. The rise detected in cfDNA HPV by ddPCR was indicative of disease progression 5 weeks before detection by CT scan and at least 10 weeks before symptomatic presentation.

### Ultra-deep cfDNA sequencing of HPV-ve OPSCC

In contrast to HPV+ve OPSCC, there are currently no generic cfDNA assays available to monitor HPV-ve OPSCC patients. Since OPSCC tumours contain characteristic genomic aberrations [30, 31], we carried out targeted ultra-deep sequencing using the Illumina TSO500 platform, on cfDNA from five HPV-ve OPSCC patients, each with matched pre- and post-treatment samples, as well as on a control DNA with spiked-in mutations, with the aim of identifying possible tumour-derived somatic mutations that could serve as markers of therapeutic response. Validation on control DNA with spiked-in mutations at 0.5% variant allele frequencies (VAF) detected 100% (12/12) of 12 mutations (median VAF 0.53%, min 0.32%, max 0.9%), supporting the accuracy of this platform in detecting the low levels of mutations present in cfDNA (Supplementary Figure S8).

Copy number variants (CNVs) have previously been reported in OPSCC tumour tissue [31, 32]. We detected CNVs in 11 of the 532 genes in the TSO500 platform in four of the five pre-treatment cfDNA samples (Figure 5A). The large majority of these CNVs were copy number losses (93.3%) with a single copy number gain noted over the BRCA2 gene in one of the pre-treatment blood samples. All patients showed a reduction in the number of CNVs following treatment, with four of the five patients exhibiting no detectable CNV post CRT (Figure 5A).

**Figure 5.**
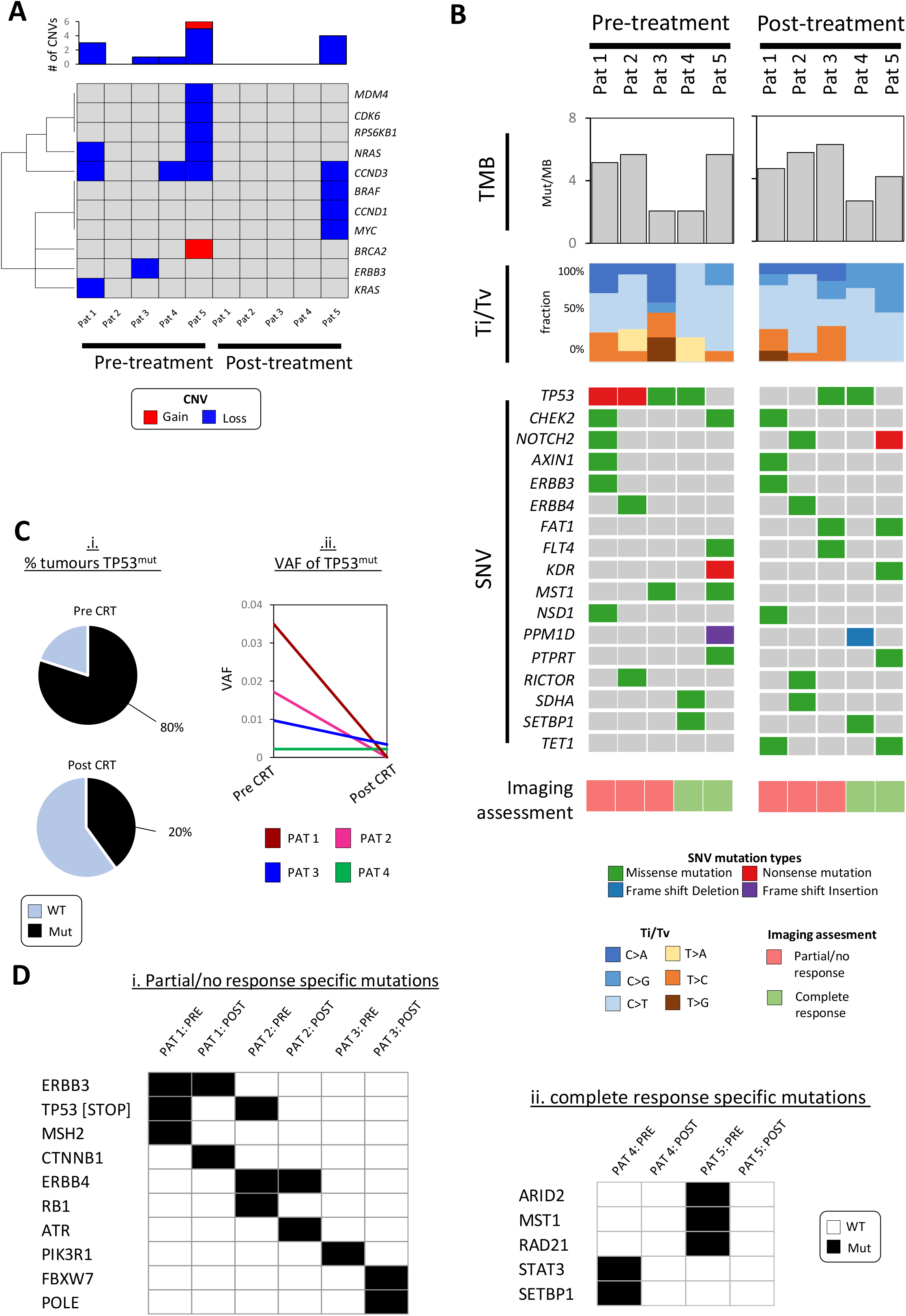
Longitudinal genomic profiling of cfDNA in HPV-ve OPSCC. **A.** Analysis of 11 genes displaying copy number variation (CNV). Blue: copy number loss, Red: copy number gain, Grey: no change. Total number of detected CNVs in each patient sample are plotted above. **B.** Molecular profiling across the 5 pre- and post-treatment HPV-ve cfDNA samples. Each column reflects a patient sample. Tumour mutational burden is plotted in the top panel (TMB) as mutations per sequenced megabase (Mut/MB). Relative percentages of transitions ad transversions (Ti/Tv) are plotted in the second panel as stacked bar charts (see the associated key). An oncoplot of single nucleotide variants (SNVs) detected in at least 20% of the samples is shown in the next panel. Mutational type is reflected by colour (see the associated key). Information on the responsiveness of each patient to treatment is shown in the bottom plot (see the associated legend). **C.** TP53 mutation in cfDNA of HPV-ve patients. (i) Pie chart of TP53 mutations in pre- (top) and post- (bottom) treatment groups. Light blue: no detected TP53 mutation, Black: TP53 mutant. (ii) Plot of change in TP53 mutation variant allele frequency (VAF) between pre- and post-treatment samples. **D.** Binary heatmap plot of mutations specific to either partial/no response (i) or complete response (ii) groups. White: no mutation detected (“WT”: wild-type), black: mutation detected (“Mut”: mutated).

In addition to CNVs, we identified a total of 83 nucleotide variants across the 10 patient samples (average: 9.5, min: 3, max: 11)(Supplementary Figure S9), detected down to a VAF of 0.09% (median 0.67%; Supplementary Figure S10). Median VAF scores did not significantly change between pre- and post-treatment samples (median pre CRT 0.71%, post CRT 0.55%. Supplementary figure S11). The large majority of the detected mutations were single nucleotide variants (SNVs; 95.2% of all variants), with a small number of INDELS also observed (1.2% insertion, 3.6% deletion)(Supplementary Figure S9). In addition, we analysed the rate of transitions and transversions in our samples which revealed an overall enrichment for C>T transitions and depletions for T>G and T>A transversions. Although we did not detect any significant difference in the tumour mutational burden pre- and post-treatment, we did detect a number of changes in the mutations between the two time points (Figure 5B & Supplementary Figures S12 & S13). Interestingly, we identified a high level of TP53 mutation in our pre-treatment cfDNA samples (4/5 patients; 80%). TP53 VAFs were greatly reduced in the matched post-treatment blood samples (Figure 5C). Other frequently mutated genes included CHEK2, NOTCH2, AXIN1, ERBB3 & ERBB4, although mutations in these genes were detected at similar levels in pre- and post-treatment samples (Figure 5B & Supplementary Figure S13).

Across the five patients, two were classified as having had a complete response and three as partial/no response following CRT. Analysis of the transitions and transversion rates across these two groups showed fewer T>A, T>C and T>G changes in individuals who responded well to CRT, both prior to as well as following treatment (Figure 5B), although given the small numbers, confirmation of this result would be needed in a larger cohort.

Analysis of differential SNVs between the complete responders compared to those with partial/no response revealed a number of mutations specific to each response group (Figure 5D). Of these, we note presence of TP53 nonsense mutations in two out of the three poor outcome HPV-ve OPSCC patients, ((Patient 1: Glu271Ter, dbSNP ID rs1060501191; Patient 2: Lys164Ter, dbSNP ID rs121912659; Supplementary Figure S14), both of which have already been classified as either associated with cancer predisposition or as pathogenic in other cancer types (https://www.ncbi.nlm.nih.gov/clinvar/). In addition, one of the TP53 missense mutations (Patient 3: Val157Phe, dbSNP ID rs121912654) has been previously reported to occur in OPSCC solid tumours [33].

## Discussion

The clinical management of OPSCC is heavily influenced by the classification into either HPV+ve or – ve tumour types [2] with clinical trials under way to investigate de-escalation of therapy in HPV+ve disease in recognition of its greater responsiveness to therapy and higher likelihood of relapse-free survival [3, 34]. Improved methods of determining and monitoring HPV status in HPV+ve disease may therefore assist in clinical decision making. Similarly, it is important to monitor tumour burden in HPV-ve patients, as these tumours present a higher risk of relapse. Current monitoring of tumour burden is reliant on imaging techniques such as CT scans and CT-FDG PET imaging. However, such tests are costly, are carried out at infrequent intervals and often provide indeterminate results. Here we benchmark the analysis of HPV in circulating cfDNA against conventional assays of tumour tissue to determine HPV status, as a guide to initial clinical decision-making. In addition, in longitudinal studies, we monitor treatment success and tumour burden in both HPV+ve and HPV-ve OPSCC by ddPCR and next-generation sequencing of plasma cfDNA.

Current assays of solid tumour to determine HPV status by p16 IHC or HPV PCR may be compromised by intra-tumour heterogeneity and reduced specificity due to the number of upstream pathways that can lead to p16 overexpression [35] [36, 37]. Accordingly, p16+ve/HPV-ve discordance rates have been reported ranging from 11-20% [37–39]. Liquid biopsy has the potential to avoid some of these difficulties by providing superior quantitation in the number of HPV genome copies compared to standard PCR assays, avoiding over-reliance on the p16 expression levels assessed by IHC and reducing inconsistencies in analysis of solid tumour biopsy due to intra-tumour heterogeneity.

In our cohort, we found a high level of concordance (93%) between ddPCR of cfDNA and p16 IHC, which was comparable to the concordance (92%) between p16 IHC and HPV PCR of tumour tissue. This value is superior to studies employing a cfDNA test against only HPV16 (81% and 71% respectively)[15, 40] – highlighting the advantage in multiple genotype based analysis in monitoring HPV+ve disease. In our study, in patients where all three assays (p16 IHC, PCR of tumour tissue, ddPCR of plasma cfDNA) were performed, 100% of samples that were positive by ddPCR were positive by either IHC or PCR whilst 85% of negative ddPCR tests were negative by either p16 IHC or PCR, consistent with other reports using various HPV analyses of cfDNA, that indicate high sensitivity and specificity of cfDNA assays for determining HPV status [11, 19]. To the best of our knowledge this is the first study comparing cfDNA ddPCR for HPV against both p16 IHC and HPV PCR on matched solid tissue samples. Taken together, the data suggest that ddPCR of cfDNA, a minimally invasive test that has low cost and rapid turnaround, has the potential to complement conventional tests of HPV status on biopsied tumour tissue.

The analysis of the general properties of cfDNA such as concentration and fragment size have previously been shown to be informative in a number of cancer types [29, 41, 42]. In contrast to a recent study, we did not observe higher concentrations of cfDNA in patients with OPSCC with respect to healthy controls [43]. We did however find that cfDNA fragment size differs between healthy controls and patients with OPSCC and that these differences reverted towards normal with treatment. cfDNA fragment size analysis may therefore add additional information that could be useful for clinical decision making, though larger studies will be required to define assay sensitivity, specificity and predictive value.

The minimally invasive nature of liquid biopsy enables repeated patient blood sampling and assay of plasma cfDNA in a way that is not easily possible with tissue biopsy. Here we demonstrate the value of longitudinal pre- and post-treatment cfDNA assessment in the clinical management of both HPV+ve and HPV-ve OPSCC. Monitoring of cfDNA HPV levels by ddPCR revealed a fall to undetectable across all patients following completion of treatment, consistent with other recent reports highlighting a correlation with reduced OPSCC tumour burden following treatment [15–17,19].

The clear fall in cfDNA HPV copy number contrasted, however, with the results of imaging. Of the 20 patients who had both CT/PET and ddPCR cfDNA within a 12 +/− 6 week window post-treatment, only 12 patients were classified as showing a complete response by imaging, whereas 18 of these patients had no detectable cfDNA HPV by ddPCR. This discrepancy may be in part due to possible overestimation of residual disease at 12 week CT/PET scans where outcomes at a later time point would suggest a value nearer 20%, or alternatively could be due to the reduced shedding of HPV DNA in low volume post-CRT disease. However, the ddPCR results are in keeping with the low recurrence rate of HPV+ve disease and the recognised high proportion of patients whose post-treatment imaging results are deemed indeterminate [44].

In our study we highlight how longitudinal monitoring by liquid biopsy has the potential to assist clinical management. In one patient (Patient #32) classified as p16 negative by IHC, the decision was made to undertake surgical intervention, which in this patient resulted in resection of tissue that did not harbour malignancy and resulted in surgical morbidity. Definition of HPV status by the cfDNA ddPCR and tissue PCR results would have deemed this patient to be HPV+ve, in which case, surgery may not have been advised. In a second patient (Patient #15) who was HPV+ve by both solid tissue and cfDNA analysis, longitudinal monitoring of cfDNA showed an initial fall to undetectable HPV copy number after CRT, and subsequently a rise in HPV copy number 5 weeks prior to the patient’s scheduled CT scan and 10 weeks before symptomatic presentation, surgery and histological confirmation of disease progression. More frequent cfDNA analyses would provide a more “real-time” assessment of tumour burden, with the potential for earlier detection of recurrence than is gained by clinical assessment or imaging, as has been observed in other cancers as well as OPSCC [11]. Together, these observations suggest that the application of ddPCR tests may help to guide clinical decision making in the management of OPSCC, particularly when test results with existing methods are discordant or indeterminate.

We also observe a single case exhibiting a much slower slow decline to undetectable HPV cfDNA levels (Patient #04). Interestingly this patient was the only HPV+ve individual who received immunotherapy raising the possibility of a difference in the kinetics of tumour ablation and/or ctDNA release in patients receiving immunotherapy. Recent work in lung adenocarcinoma studies suggests that cfDNA analysis can aid in the interpretation of imaging results: distinguishing progression from so called pseudo-progression brought upon through inflammation at the site of the tumour during anti-PD-1 treatment [45]. Further studies investigating the implications of this in larger patient cohorts would be valuable in furthering understanding of the suitability of immunotherapy in OPSCC.

Finally, unlike HPV+ve OPSCC, there is no single blood-based assay to follow OPSCC patients with HPV-ve disease. Since HPV-ve OPSCC tumours have been reported to contain discrete sets of somatic mutations [30, 31, 46], we set out to identify and monitor tumour-derived somatic mutations in cfDNA from patients with HPV-ve OPSCC. In contrast to exome sequencing from solid tumour material, sequencing of cfDNA presents challenges, owing to the low levels of tumour-derived DNA within circulating cfDNA, the majority of which is non-tumour-derived DNA. To overcome these challenges, we applied a targeted ultra-deep sequencing approach that includes wet lab and informatic features such as unique molecular identifiers and error suppression to remove duplicate reads and minimise sequencing errors without losing the signal of very low frequency (<1%) sequence variations. To improve further on small variant calling results, bioinformatic pipelines performed local indel realignment, paired read stitching (i.e. a single read that has been combined from a pair of reads) and read filtering based on QC metrics. The net effect of the read collapsing and TMB analysis steps reduces false positives in a typical cell-free DNA sample from ∼1500 per Mb to < 5 per Mb. The approach therefore offers an improvement over alternative panel-based sequencing strategies through the reduction of errors that are mostly of a technical nature.

The sequencing of control DNA with spiked-in mutations revealed a high degree of sensitivity with all mutations (12/12) detected and accurate to within 0.4% in detecting mutations at 0.5% VAF. In this work we generate, to our knowledge, the first genomic signatures of ctDNA in HPV-ve OPSCC, comprising copy number, mutational signature and single nucleotide changes down to 0.09% (ARID2 frameshift mutation in patient #5). Our data detected CNVs in the cfDNA of four of the five HPV-ve patients analysed, with a reduction in CNVs following treatment: four of the five patients exhibited no CNVs following CRT. Although we did not observe some of the CNVs previously reported in OPSCC tumours, possibly due in part to the small cohort size of our sequenced dataset, we found a number of single nucleotide mutations in the TP53 gene and in members of the ERBB family in keeping with previous OPSCC tumour data [31, 32]. We also identified candidate mutations in cfDNA that are of potential prognostic value, such as the observation that nonsense TP53 mutations in our cohort were found exclusively in individuals with poor response to CRT, a result that it would be of interest if confirmed in a larger cohort of pre- and post-treatment OPSCC patients. Interestingly this is in agreement with published reports that state the presence of mutations in TP53 were found associated with reduced recurrence-free survival in patients with OPSCC (42.6% TP53 mutant versus 75.9% TP53 WT relapse rate across a median 26 month follow up study)[46].

It is anticipated that mutation load would reduce upon response to CRT although clonal selection and outgrowth of previously undetected mutations could occur post-treatment [47]. In our patients, we found 2/5 cases that show a reduction, 2/5 cases an increase and 1/5 no change in mutation count following treatment, with no correlation to treatment response as assessed by imaging. We did however observe a small reduction in the overall VAFs of mutated genes in post-CRT samples suggesting the possibility that identifiable mutation frequencies decrease after completion of treatment. Further assessment of this ultra-deep sequencing method in larger cohorts will allow for more accurate assessment of the technical and/or biological factors affecting detection of tumour-derived mutations in cfDNA.

Taken together, this study combines novel cfDNA findings to complement and in some cases enhance current methods to characterise and monitor both HPV+ve and HPV-ve OPSCC patients throughout their clinical care. Expansion of these findings into larger cohorts with further long-term follow-up will help to establish more definitively the role of cfDNA analysis in personalised management of OPSCC.

## Data Availability

The primary and processed data generated on the TSO500 platform are currently being submitted to the European Genome-phenome Archive (EGA; https://ega-archive.org/access/data-access). The data will be made available upon request to the authors.

## Acknowledgements

We extend our thanks to the patients who contributed to this study. We thank Edinburgh Genomics, Edinburgh, UK for their support with high throughput sequencing. We thank the NRS Lothian Human Annotated Bioresource, NHS Lothian Department of Pathology, the Scottish HPV Reference Laboratory, Edinburgh Experimental Cancer Medicine Centre and NHS Lothian Clinical Genetics for clinical and laboratory support. We thank Illumina for early access to the TSO500 platform, provision of sequencing kits and for technical and bioinformatic support. We thank Lara Carey and Peter G Morrice for laboratory assistance and Sophie Marion De Proce, Robert L Hollis and Simon C Herrington for comments on this manuscript.

